# A comparison of Emergency Department presentations for Medically Unexplained Symptoms in Frequent Attenders during COVID-19

**DOI:** 10.1101/2020.08.25.20181511

**Authors:** Natasha Faye Daniels, Raiiq Ridwan, Ed B G Barnard, Talha Muneer Amanullah, Catherine Hayhurst

## Abstract

**Background:** Medically Unexplained Symptoms (MUS) refer to symptoms with no identified organic aetiology, and are amongst the most challenging for patients and Emergency Department (ED) staff. Providers working in our ED perceived an increase in severity and frequency of these types of presentations during the COVID-19 pandemic.

**Methods:** A retrospective list of frequent attenders (FA) presenting five or more times to the ED between two 122-day periods were examined: 01 Mar to 30 Jun 2019 (Control) and 2020 (COVID-19). The FA group were then examined to identify patients presenting with MUS (FA-MUS). Data were analysed in Prism; presented as n(%), % (95% confidence interval (95%CI) – Wilson/Brown method). Proportions were compared with a two-tailed Fisher’s exact test. A Baptista-Pike odds ratio was used to estimate magnitude and precision.

**Results:** The total number of ED attendances during the control period was n=42,785 which reduced to n=28,806 in the COVID-19 period, a decrease of 32.7%. The control FA cohort had *n*=44 FA-MUS patients with 149 ED visits. This increased to *n*=65 FA-MUS patients with 267 visits during COVID-19, *p*=0.44. There was a significant increase in the proportion of all ED visits that were FA-MUS: 0.3% (control) compared to 0.9% (COVID-19); OR 2.7, *p*<0.001. There was a significant increase in shortness of breath amongst MUS during the COVID-19 pandemic relative to the control period (p<0.01), with no significant difference in any other MUS category.

**Conclusion:** Whilst the total number of ED attendances reduced by almost one third during COVID-19, the actual number of all visits by frequent attenders with MUS increased and the proportion of attendances by these tripled during the same period. This presents an increasing challenge to ED clinicians who may feel underprepared to manage these patients effectively.

What is already known
1. Medically Unexplained Symptoms (MUS) are those that have no identified organic aetiology - they are amongst the most challenging presentations for patients and Emergency Department (ED) staff.
2. During times of stress and uncertainty, frequent attenders (FA) appear to be disproportionately affected by MUS. However, there are few data examining the impact of COVID-19 on the FA population.

What this paper adds
1. There was a significant increase in the proportion of all ED visits by FAs during the first four months of the COVID-19 pandemic.
2. There was a significant increase in the proportion of all ED visits by FAs with MUS during the first four months of the COVID-19 pandemic.
3. The proportion of MUS presentations that were ‘shortness of breath’ was significantly higher in the COVID-19 period compared to the control period. There were no other proportional differences observed in MUS categories.

## Introduction

Since the declaration of the COVID-19 pandemic, the physical and psychological sequelae associated with SARS-CoV2 are still being discovered. The pandemic’s impact on mental health has been extensively discussed in the literature, with COVID-19 related health anxiety admissions described.[1] Particularly challenging ED presentations are those in which symptoms have no identified organic aetiology, referred to as Medically Unexplained Symptoms (MUS).[2] These symptoms include non-cardiac chest pain, gastrointestinal complaints, non-epileptic seizures, functional neurology, and shortness of breath.

MUS is a common presentation in frequent attenders (FA). In a previously published dataset from our hospital, 45% of FA were identified as having one or more MUS.[3] The Royal College of Emergency Medicine defines FA’s as those who attend an ED five times or more in a year. The impact of MUS on patients can be debilitating, with added stressors due to stigma experienced both within society and the healthcare system.[4] Despite this, there are few data reporting rates of MUS or treatment in the ED. This issue is key for two reasons: the personal burden for patients, and the disproportionate use of allocated resources by the FA group. [5,6]

Our primary aim was to compare the proportion of FA with MUS amongst all ED attendances during the first four months of the COVID-19 pandemic compared to a control period in 2019. The secondary aim was to compare the relative frequency of MUS diagnostic categories between the two periods.

## Methods

A retrospective list of FA presenting five or more times to the ED at Cambridge University Hospitals between two 122-day periods were examined: 01 Mar to 30 Jun 2019 (Control) and 2020 (COVID-19). The hospital is a Major Trauma Centre and tertiary referral centre with an annual ED census >120,000.

Examining case notes of FAs, we categorized each presentation as MUS, physical health or mental health visits (overdoses, deliberate self-harm, episodes of psychosis etc.). The MUS presentations were included in the study for both the COVID-19 and Control groups. MUS presentations were subsequently categorized into seven categories: non-cardiac chest pain, abdominal pain, functional neurological symptoms, non-epileptiform seizures, musculoskeletal symptoms, shortness of breath, and other.

### Data analysis

Data were analysed in Prism for macOS (v.8.4.2, GraphPad Software, San Diego, CA, USA); presented as n(%), % (95% confidence interval (95%CI) – Wilson/Brown method). Proportions were compared with a two-tailed Fisher’s exact test. A Baptista-Pike odds ratio (OR) with 95%CIs was used to estimate magnitude and precision.

## Result

### Overall activity

There were a total of *n*=42,785 ED visits in the control period and *n*=28,806 in the COVID-19 period, a decrease of 32.7% (95%CI 32.2-33.1).

### Frequent attender visits

During the control period, there were *n*=163 FA patients with 1185 ED visits (7.3 visits/pt), compared to *n*=147 patients with 1000 visits (6.8 visits/patient), *p*=0.58. The proportion of all ED visits by FA was significantly higher in the COVID-19 period: 3.5% (COVID-19) compared to 2.8% (control), OR 1.3 (95%CI 1.2-1.4), *p*<0.001.

### MUS visits

There were *n*=44 FA-MUS patients with 149 ED visits in the control period, and *n*=65 with 267 visits in the COVID-19 period; 3.4 versus 4.1 visits/patient, *p*=0.44. There was a significant increase in the proportion of all ED visits that were FA-MUS: 0.3% (control) compared to 0.9% (COVID-19); OR 2.7 (95%CI 2.2-3.3), *p*<0.001.

### MUS categories

FA-MUS visits were categorised into common syndromes; categories with less than ten total visits were grouped into ‘other’ (for example, palpitation (*n*=9), falls (*n*=6), vomiting (*n*=5)). The only difference observed was a significant increase in the MUS diagnosis of shortness of breath during the COVID-19 period, Table 1.

**Table 1:**
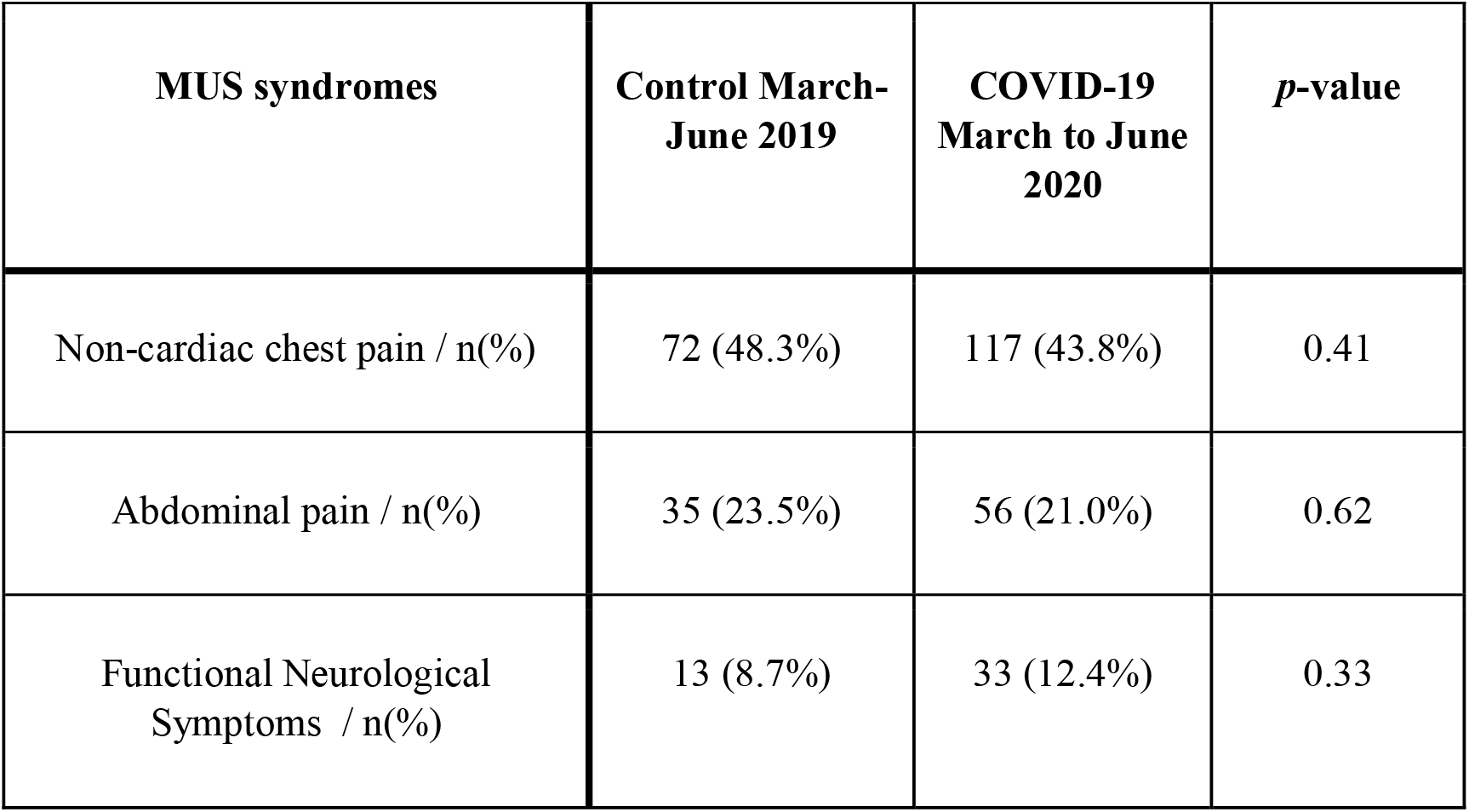
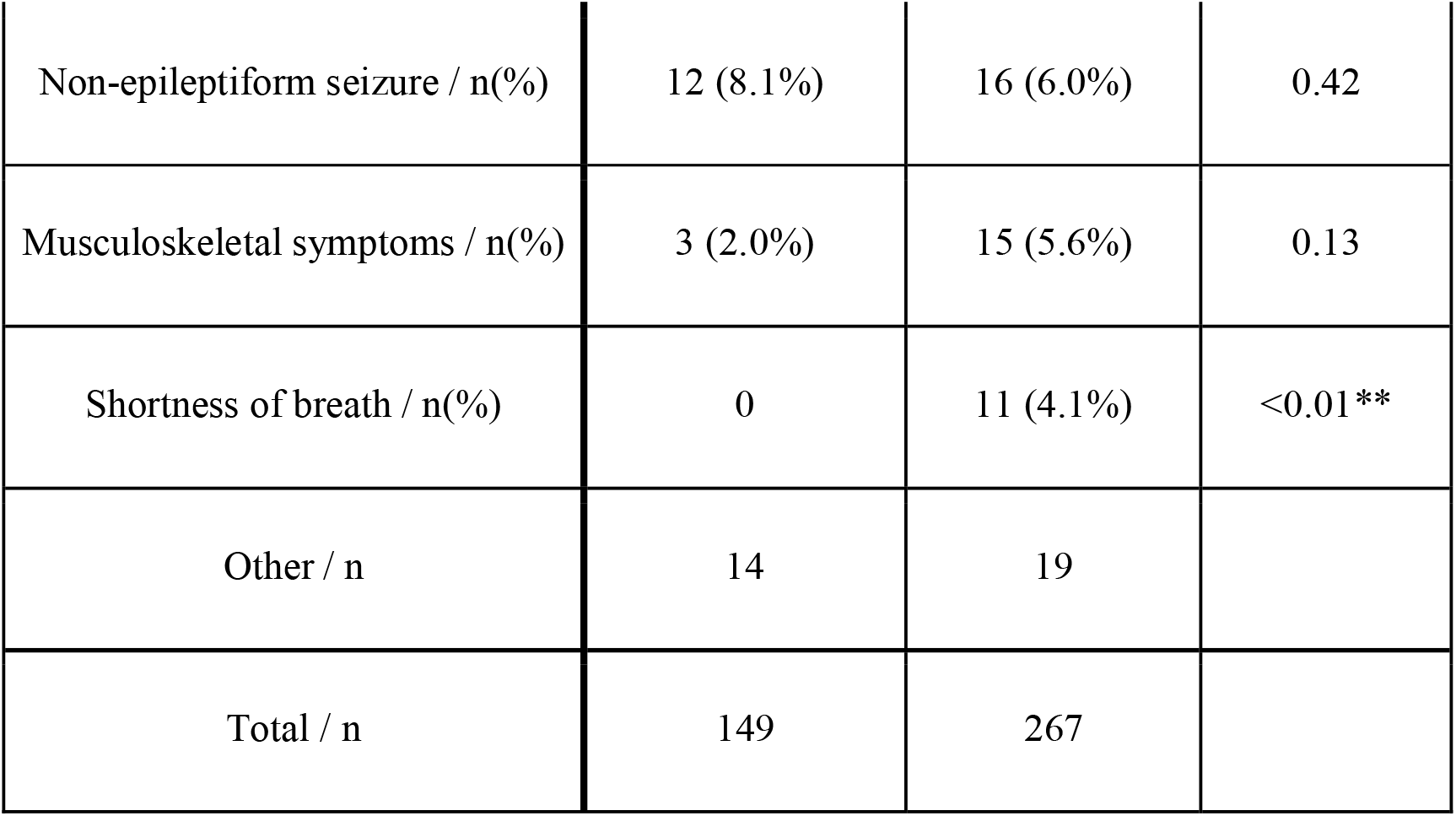
Categories of MUS presenting features.

| MUS syndromes | Control March-June 2019 | COVID-19 March to June 2020 | *p*-value |
| --- | --- | --- | --- |
| Non-cardiac chest pain / n(%) | 72 (48.3%) | 117 (43.8%) | 0.41 |
| Abdominal pain / n(%) | 35 (23.5%) | 56 (21.0%) | 0.62 |
| Functional Neurological Symptoms / n(%) | 13 (8.7%) | 33 (12.4%) | 0.33 |
| Non-epileptiform seizure / n(%) | 12 (8.1%) | 16 (6.0%) | 0.42 |
| Musculoskeletal symptoms / n(%) | 3 (2.0%) | 15 (5.6%) | 0.13 |
| Shortness of breath / n(%) | 0 | 11 (4.1%) | <0.01** |
| Other / n | 14 | 19 |  |
| Total / n | 149 | 267 |  |

## Discussion

This study has demonstrated a significant increase in FA-MUS attendances to our ED during COVID-19, a phenomenon seen in other cases of unpredictable threats.[7] This suggests that despite the risk of attending an ED in the context of a pandemic, these symptoms are debilitating enough for the patients to deem the risk necessary. Shortness of breath was the only MUS presentation that significantly increased during COVID-19, likely secondary to pandemic related anxiety.

Despite the difficulties in managing MUS patients, to the best of our knowledge, there are no studies looking at how the COVID-19 pandemic has influenced this population. Generally the default management of the MUS patient is extensive investigation to rule out physical aetiology, followed by psychiatric assessment and/or discharge.[8] However, it has been reported that early diagnosis with reassurance and an explanation regarding the mechanisms of such symptoms can help.[4] Alternatively, a stepped Psychological approach may be of benefit.[9]

ED providers often report uncertainty in managing patients with MUS, prompting a need for increased knowledge so that investigations to rule out other pathology are balanced with early diagnoses and appropriate interventions. Some of these patients would be better served within primary care but this depends on ease of access and primary care clinicians being confident of MUS diagnosis and management.

The high prevalence of FA in the ED is likely a symptom of the general trend of unmet needs for this diverse and vulnerable group elsewhere in the healthcare system.[10] This paper adds further evidence that the needs of these individuals with MUS are not being met, and in added stressors such as the COVID-19 pandemic, their needs are further exacerbated.

### Limitations

The data analysis was performed retrospectively and represents a single ED, thus the results are not necessarily translatable to other centres. Additionally, due to the perceived increase in MUS patients before commencing the data extraction, reviewer bias is an important consideration. This was minimised by having two reviewers (ND and RR) extract the data from both 2019 and 2020.

## Conclusion

Whilst the total number of ED attendances reduced by almost one third during COVID-19, the proportion of all visits by FA-MUS tripled during the same period. This paper highlights the significance of the MUS experience, with patients willing to risk their safety at the peak of the pandemic. This speaks volumes of the severity of the FA-MUS patient experience, and should prompt the general healthcare system to consider how to better help this patient group.

## Data Availability

Anonymised data are available from the primary author at reasonable request

## Contribution statement

The study was conceived by CH. Data acquisition was performed by ND and RR. Data analysis was performed by EB and TA. The manuscript was drafted by ND and EB. Revisions were performed by CH and EB. All authors have contributed to and agreed the final version.

## Acknowledgements

We thank Dr Shiraz Hashmi (Aga Khan University Hospital) for his help in organizing the data and giving statistical input.

## Patient and Public Involvement

No patient or public involvement.

